# Development and Prospective Implementation of a Large Language Model based System for Early Sepsis Prediction

**DOI:** 10.1101/2025.03.07.25323589

**Authors:** Supreeth P. Shashikumar, Sina Mohammadi, Rishivardhan Krishnamoorthy, Avi Patel, Gabriel Wardi, Joseph C. Ahn, Karandeep Singh, Eliah Aronoff-Spencer, Shamim Nemati

**Affiliations:** Division of Biomedical Informatics, UC San Diego, San Diego, USA; Department of Emergency Medicine, UC San Diego, San Diego, USA; Division of Pulmonary, Critical Care and Sleep Medicine, UC San Diego, San Diego, USA; Division of Gastroenterology and Hepatology, Mayo Clinic, Rochester, USA; Jacobs Center for Health Innovation, UC San Diego Health, San Diego, USA; Division of Infectious Diseases and Global Public Health, UC San Diego, San Diego, USA

## Abstract

Sepsis is a dysregulated host response to infection with high mortality and morbidity. Early detection and intervention have been shown to improve patient outcomes, but existing computational models relying on structured electronic health record data often miss contextual information from unstructured clinical notes. This study introduces COMPOSER-LLM, an open-source large language model (LLM) integrated with the COMPOSER model to enhance early sepsis prediction. For high-uncertainty predictions, the LLM extracts additional context to assess sepsis-mimics, improving accuracy. Evaluated on 2,500 patient encounters, COMPOSER-LLM achieved a sensitivity of 72.1%, positive predictive value of 52.9%, F-1 score of 61.0%, and 0.0087 false alarms per patient hour, outperforming the standalone COMPOSER model. Prospective validation yielded similar results. Manual chart review found 62% of false positives had bacterial infections, demonstrating potential clinical utility. Our findings suggest that integrating LLMs with traditional models can enhance predictive performance by leveraging unstructured data, representing a significant advance in healthcare analytics.

## Introduction

Sepsis is a dysregulated host response to infection that is estimated to affect over 48 million adults worldwide every year, resulting in approximately 11 million deaths^1,2^. In the United States alone, one in three hospital deaths is attributable to sepsis^3^. Studies have shown that early recognition of sepsis followed by timely interventions such as appropriate antibiotic administration, fluid resuscitation, and source control can lead to improved patient outcomes^4–10^. The high mortality rate and substantial economic burden of sepsis, estimated at over $32,000 per patient in high-income countries^11^, underscore the urgent need for effective early detection and intervention strategies. Consequently, extensive efforts have been made to develop tools for the early prediction of sepsis using electronic health record (EHR) data^12^. Additionally, a limited number of studies have demonstrated improvements in patient outcomes following the real-world deployment of sepsis prediction models^13–15^.

Although EHRs contain both structured data (vital signs, laboratory results, comorbidities, etc.) and unstructured data (triage notes, progress notes, radiology reports, etc.), most sepsis prediction models have primarily utilized structured data^12^. Unstructured clinical data, including triage notes, provider notes, and progress notes, often contain valuable contextual information that structured data cannot capture. For instance, summaries of clinical signs and symptoms at hospital presentation, initial physical exams, and interpretations of radiological exams can be crucial for diagnosing or ruling out sepsis. Hence, incorporating unstructured data could enhance the performance of sepsis prediction models^16–20^. A common approach to using unstructured data has been obtaining document-level representations (i.e., embeddings) and using them as inputs to the sepsis prediction model alongside structured data. These document-level representations can be obtained using pre-trained language models (such as ClinicalBERT)^16,17,19,21,22^, topic modeling techniques (such as LDA)^18,23^, or term frequency-inverse document frequency (tf-idf) embeddings based on clinical entities extracted from clinical notes^18,19,22,24^. However, these approaches have several limitations^25,26^: 1) they capture information at a document level (or global level) and may not effectively capture local contextual information relevant to sepsis; 2) they offer limited interpretability; 3) may overly emphasize repeated information due to redundant text from copy-and-paste practices^27^ ; and 4) they are prone to domain shift, as such approaches may capture hospital-specific data entry patterns. These limitations could potentially be addressed with the use of large language models (LLMs) and techniques such as retrieval augmented generation (RAG)^28^.

LLMs are foundation models built with billions or trillions of parameters, trained on extensive text data to recognize complex language patterns^29^. Recently, LLMs have garnered significant attention in the field of medicine for their capability to use natural language to generate coherent, contextually relevant responses. Initial investigations have demonstrated successful applications of LLMs in identifying social determinants of health^30^, assigning diagnosis-related groups^31^, answering medical questions^32^, drafting clinical notes^33^, and drafting clinician responses to patient inquiries^34,35^. Of note, LLMs have proven effective in retrieving and processing relevant information from multiple documents^36^.

The goal of this study was to develop and prospectively validate a multimodal system (COMPOSER-LLM) that combines LLM-based processing of unstructured clinical notes with structured data from the EHR to enhance model accuracy in challenging diagnostic scenarios. We hypothesized that COMPOSER-LLM’s incorporation of clinical notes for differential diagnosis of sepsis-mimics would outperform our previous binary sepsis prediction model (COMPOSER) that relied solely on structured EHR data. Please see the subsection *“Differential diagnosis tool”* in Methods for a list of all the sepsis-mimics. The entire pipeline was hosted on a cloud-based healthcare analytics platform that used Fast Healthcare Interoperability Resources (FHIR) and Health Level Seven International Version 2 (HL7v2) standards to provide real-time access to data elements. This allowed us to evaluate the model’s performance in a real-world setting, where clinical notes may be incomplete. The schematic diagram of the entire COMPOSER-LLM pipeline is shown in Figure 1. Please note that throughout this paper, COMPOSER-LLM and COMPOSER-LLM*_DDx_* refer to the same pipeline, unless otherwise noted.

**Figure 1.**
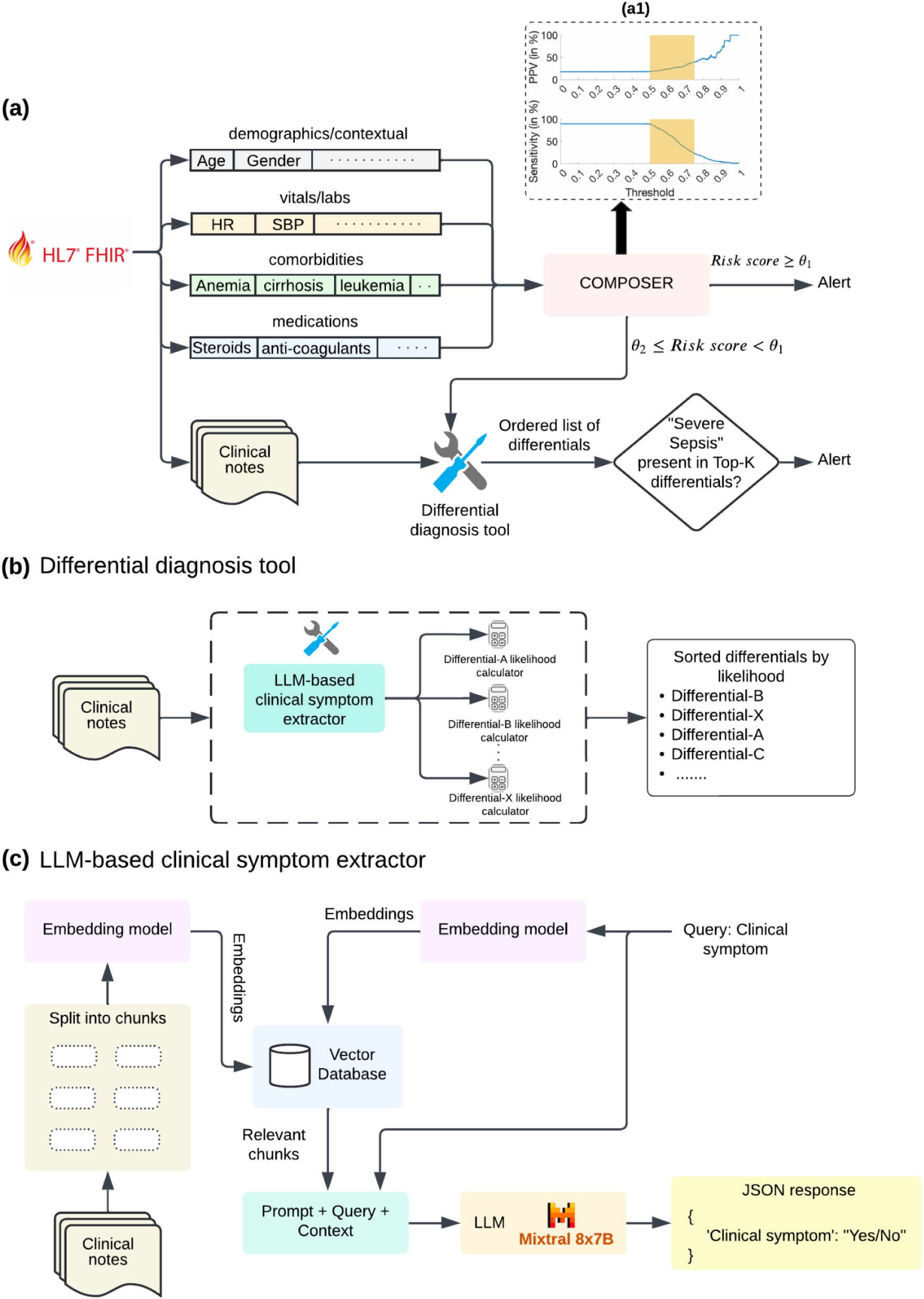
Schematic Diagram of the COMPOSER-LLM pipeline. **(a)** The entire COMPOSER-LLM pipeline was hosted on a cloud-based healthcare analytics platform that used FHIR and HL7v2 standards to provide real-time access to data elements. The input feature set (including laboratory measurements, vitals measurements, comorbidities, medications and demographics) was queried using FHIR APIs and passed to the COMPOSER model. For samples with a sepsis risk score greater than a primary decision threshold (θ_1_), an alarm was fired based on the standalone composer risk score. However, for samples with a sepsis risk score falling within a high uncertainty score region (θ_2_ ≤ sepsis risk scores <θ_1_), the LLM-based differential diagnosis tool was utilized to improve diagnostic certainty. The high uncertainty score region is shown as the shaded region in the inset figure. Finally, if ‘Severe Sepsis’ was present within the top-5 sorted differentials and the LLM identified a ‘suspicion of bacterial infection’, an alarm was fired. **(b)** The differential diagnosis tool employed an LLM to extract severe sepsis and sepsis-mimic (or differentials) related clinical signs and symptoms from the clinical notes. These identified symptoms were then fed into likelihood calculators to determine the likelihood of various differentials. The resulting differential diagnosis list was sorted by the likelihood scores. **(c)** The LLM-based clinical sign or symptom extractor was designed to take a prompt (containing the clinical sign or symptom to be queried) and clinical notes (all notes generated from admission to the prediction time) as input and produce output in a JSON format.

## Results

### Study population

During the study period for retrospective model development (September 1, 2023 through December 31, 2023), a total of 1,746 emergency department (ED) encounters (16.6% septic) met the inclusion criteria (*retrospective cohort*) for this study. Separately, the study period for prospective validation of the model included May 1, 2024 through June 15, 2024. During this period, a total of 754 ED encounters (18.4% septic) met the inclusion criteria for this study (*prospective cohort)*. Table 1 shows baseline characteristics and summary characteristics for the *retrospective* and *prospective* cohorts.

**Table 1:** Patient characteristics of the *retrospective* and *prospective* cohorts.

|  | Retrospective cohort |  | Prospective cohort |  |
| --- | --- | --- | --- | --- |
|  | *Septic* | *Non-Septic* | *Septic* | *Non-Septic* |
| # Encounters (%) | 290 (16.6%) | 1456 | 139 (18.4%) | 615 |
| Age (in years), median [IQR] | 64.5 [51.4 - 75.1] | 59.1 [44.2 - 71.1] * | 65.1 [50.1 - 75.8] | 58.1 [43.3 - 69.6] * |
| Gender (Male), % | 54.1% | 51.5% | 58.2% | 47.7% |
| Race |  |  |  |  |
| White, % | 45.1% | 44.4% | 50.4% | 45.6% |
| African American, % | 10% | 9.4% | 7.2% | 7.9% |
| Asian, % | 7.2% | 6.8% | 4.3% | 6.4% |
| ED Length of Stay (in hours), median [IQR] | 24.9 [11.2 - 50.6] | 11.3 [6.5 - 28.3] * | 22.1 [10.5 - 45.8] | 11.9 [6.9 - 30.8] * |
| CCI, median [IQR] | 2 [0 - 4] | 1 [0 - 3] | 2 [0 - 5] | 1 [0 - 2] |
| SOFA, median [IQR] | 3 [1 - 5] | 1 [0 - 2] * | 3 [1 - 5] | 1 [0 - 2] * |
| In-hospital mortality, % | 8.9% | 1.5%* | 8.1% | 1.2% * |
| Time from ED triage to onset of sepsis (in hours), | 3.2 [1.6 - 7.5] | N/A | 2.3 [1.2 - 10.2] | N/A |

\* p-value<0.05
CCI = Charlson Comorbidity Index
SOFA = Sequential Organ Failure Assessment score

### Performance of Composer-LLM_DDx_ pipeline

The standalone COMPOSER model achieved a sensitivity of 72.9%, a positive predictive value (PPV) of 22.6%, F-1 score of 34.5%, and false alarms per patient hour (FAPH) of 0.037 (e.g., 1.48 false alarms every 2h in a 20-bed care unit) on the retrospective validation cohort. When the LLM-based differential diagnosis tool was applied to COMPOSER risk scores in the range of 0.5 to 0.75 (θ_2_ = 0.5, θ_1_ = 0.75), COMPOSER-LLM*_DDx_* demonstrated improved performance with a sensitivity of 72.1%, a PPV of 52.9%, F-1 score of 61.0%, and FAPH of 0.0087 (e.g., 0.348 false alarms every 2h in a 20-bed care unit) on the same cohort. In comparison, when the LLM-based sepsis likelihood tool alone was applied to COMPOSER risk scores in the range of 0.5 to 0.75 (θ_2_ = 0.5, θ_1_ = 0.75), COMPOSER-LLM*_SLT_* demonstrated a lower performance with a sensitivity of 72.1%, a PPV of 31.9%, F-1 score of 44.2%, and FAPH of 0.021 (e.g., 0.84 false alarms every 2h in a 20-bed care unit) on the same cohort. Performance metrics of all models on the retrospective development cohort are detailed in Supplementary Table 1. Note that, for COMPOSER risk scores greater than the primary decision threshold (θ_1_), an alert was fired irrespective of the output from LLM. The performance of the COMPOSER-LLM*_DDx_* pipeline for running the LLM-based sepsis likelihood tool for various risk score intervals is shown in Supplementary Table 3. Additionally, outputs generated by the LLM (for clinical signs or symptoms related to severe sepsis) for a sample patient are shown in Supplementary Table 2.

Finally, COMPOSER-LLM*_DDx_* demonstrated superior performance compared to the COMPOSER-LLM*_baseline_* model (detailed in the Methods section under “Baseline model comparison”) which achieved a sensitivity of 34.4%, PPV of 35.1% and F1-score of 34.7% on the retrospective validation cohort. A comparison of performance of all the models considered in this study is shown in Table 2.

**Table 2:** Comparison of model performance.

|  |  | COMPOSER<sup>1</sup> | COMPOSER-LLM<sub>SLT</sub><sup>2</sup><br>(with sepsis likelihood tool) | COMPOSER-LLM<sub>DDx</sub><sup>3</sup><br>(with differential diagnosis tool) |
| --- | --- | --- | --- | --- |
| Retrospective validation cohort | Sensitivity | 72.9% | 72.1% | 72.1% |
|  | PPV<sup>4</sup> | 22.6% | 31.9% | 52.9% |
|  | F1-Score | 34.5% | 44.2% | 61.0% |
|  | FAPH<sup>4</sup> | 0.037 | 0.021 | 0.0087 |
| Prospective cohort | Sensitivity | 70.8% | 70.5% | 70.8% |
|  | PPV<sup>4</sup> | 25.1% | 36.3% | 58.2% |
|  | F1-Score | 37.1% | 47.9% | 63.9% |
|  | FAPH<sup>4</sup> | 0.034 | 0.020 | 0.0086 |

<sup>1</sup> Decision threshold of COMPOSER was set to 0.6 as previously described in Boussina et al.
<sup>2</sup> LLM-based sepsis likelihood tool was applied to COMPOSER risk scores in the range of 0.5 to 0.75 ( $\theta_2 = 0.5$ , $\theta_1 = 0.75$ )
<sup>3</sup> LLM-based differential diagnosis tool was applied to COMPOSER risk scores in the range of 0.5 to 0.75 ( $\theta_2 = 0.5$ , $\theta_1 = 0.75$ )
<sup>4</sup> PPV = positive predictive value, FAPH = false alarms per patient hour

In an additional exploratory analysis, we measured the proportion of the false positives (produced by COMPOSER-LLM*_DDx_*) that contained the actual patient diagnosis within the Top-5 differentials produced by the differential diagnosis tool. It was observed that for 83.1% of the false positive patients, the actual patient diagnosis was contained in the predicted differential diagnosis list.

### Prospective performance of COMPOSER-LLM

The COMPOSER-LLM pipeline was prospectively deployed across two EDs within the UCSD Health system starting on May 1, 2024 (see Figure 2 for an overview of the real-time deployment pipeline). The *prospective cohort* analyzed in this study included patients admitted to the ED between May 1, 2024, and June 15, 2024. In the *prospective cohort*, the COMPOSER-LLM*_DDx_* pipeline (with θ_2_ = 0.5, θ_1_ = 0.75) achieved a sensitivity of 70.8%, a PPV of 58.2%, F-1 score of 63.9% and FAPH of 0.0086 (e.g., 0.344 false alarms every 2h in a 20-bed care unit). The performance in the prospective cohort was found to be similar to that of the retrospective validation cohort, which had a sensitivity of 72.1%, a PPV of 52.9%, F-1 score of 61.0%, and FAPH of 0.0087 (e.g., 0.348 false alarms every 2h in a 20-bed care unit).

**Figure 2.**
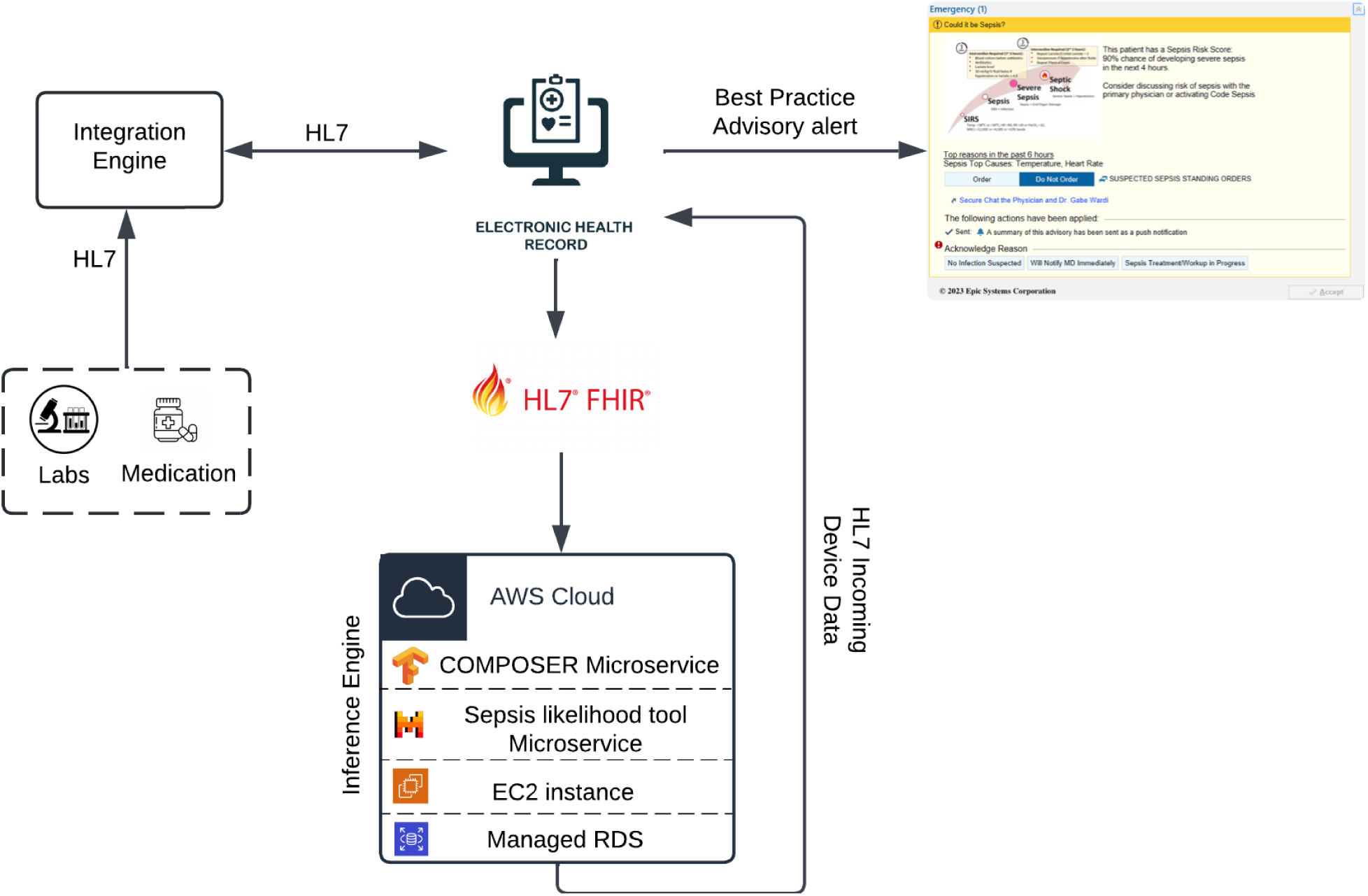
Schematic diagram of the COMPOSER-LLM real-time deployment pipeline. The real-time platform extracts data at an hourly resolution of all active patients using FHIR APIs, and passes the input feature set (consisting of laboratory measurements, vitals measurements, comorbidities, medications, clinical notes, and demographics) to the COMPOSER-LLM inference engine. The sepsis risk scores generated by the COMPOSER-LLM pipeline are then written back to the EHR (as a flowsheet item) through an HL7 device data interface. The flowsheet then triggers a nurse-facing Best Practice Advisory that alerts the caregiver that the patient is at risk of developing severe sepsis. HL7 = health level 7; AWS = Amazon Web Services; FHIR = Fast Healthcare Interoperability Resources; RDS = relational database service; EC2 = Elastic Compute Cloud

In an additional exploratory analysis, we calculated the proportion of the false positives (produced by COMPOSER-LLM*_DDx_*) that contained the actual patient diagnosis within the Top-5 differentials produced by the differential diagnosis tool. It was observed that for 73.2% of the false positive patients, the actual patient diagnosis was contained in the predicted differential diagnosis list.

### Clinician chart review of False-Positives

Chart review of 50 false-positive patients were performed by two clinicians. They were asked to examine each patient’s EHR record and determine if there was a suspicion of bacterial infection at the time the COMPOSER-LLM alarm was triggered. Clinician 1 identified 31 out of 50 patients (62%) as having a suspicion of bacterial infection, while Clinician 2 identified 32 out of 50 patients (64%). Both clinicians agreed on 31 out of the 50 false-positive patients (62%) having a suspicion of bacterial infection. The chart review indicated that the majority of patients in the false-positive group would benefit from the COMPOSER-LLM alert.

### Performance of COMPOSER-LLM for patients with clinical suspicion of infection

The goal of this sub-analysis was to explore the potential of utilizing COMPOSER-LLM*_DDx_* as a digital sepsis biomarker. Much of the current sepsis biomarkers^37^ are designed to be used for patients with a suspicion of clinical infection. Replicating a similar condition for use for COMPOSER-LLM*_DDx_*, we evaluated its performance for patients with a clinical suspicion of infection (defined by the clinical decision to order blood culture and antibiotics within a 6-hour window) at the time of prediction. Under the above condition for use, the standalone COMPOSER achieved a sensitivity of 75%, a PPV of 55.4% and F-1 score of 63.7% whereas COMPOSER-LLM*_DDx_* achieved a higher performance with a sensitivity of 74.5%, a PPV of 80.1% and F-1 score of 77.2% on the retrospective validation cohort.

A similar pattern was observed on the prospective evaluation cohort, with the standalone COMPOSER achieving a sensitivity of 70.9%, a PPV of 58.4% and F-1 score of 64.1% and COMPOSER-LLM*_DDx_* achieving a higher performance with a sensitivity of 74.4%, a PPV of 81.3% and F-1 score of 77.7% on the retrospective validation cohort.

## Discussion

We introduce a novel approach for accurate early prediction of sepsis, leveraging a locally-deployed LLM to analyze unstructured clinical notes in real-time to assess the likelihood of sepsis and sepsis-mimics. Our findings are important because they demonstrate LLMs can enhance the performance of existing clinical predictive models, particularly in high-uncertainty prediction scenarios, by incorporating and analyzing the rich clinical context found within unstructured medical notes. The entire pipeline was deployed on a cloud-based healthcare analytics platform to ensure scalability and portability, utilizing FHIR and HL7v2 standards for interoperability. The LLM-based differential diagnosis tool was employed only for patients with COMPOSER risk scores within a high uncertainty interval (near the model decision threshold). This approach enhanced prediction certainty by reducing false alarms while avoiding the continuous computational cost of running an LLM. When prospectively evaluated, the COMPOSER-LLM pipeline showed similar performance to the retrospective validation cohort; demonstrating the utility of the LLM-augmented system in the context of potentially incomplete clinical notes.

Recent work has highlighted the potential of artificial intelligence (AI) models for predicting sepsis using EHR data. These models, including neural networks, can process complex, high-dimensional data and capture nonlinear interactions between variables, demonstrating promising results in early sepsis detection. A systematic review by Islam et al. identified at least 42 studies that developed machine learning (ML) models using vital signs, laboratory data, and demographic information^12^. Many of these studies have primarily relied on structured EHR data, overlooking the rich information contained in unstructured clinical notes which may augment predictive abilities^38^. Importantly, recent data have highlighted suboptimal performance of commercially available sepsis predictive models that rely on structured data, such as the Epic Sepsis score^39,40^ (sensitivity of 33% and PPV of 12%), and TREWS^41^ (sensitivity of 71% and PPV of 16.7% for patients not already on antibiotics). In comparison, COMPOSER-LLM demonstrated superior performance with a sensitivity of 72.1% and PPV of 52.9%.

In this study, we examined the effectiveness of LLMs as information retrieval tools rather than diagnostic tools, followed by probabilistic reasoning to aid in sepsis prediction under high-uncertainty scenarios. This integrated strategy aligns with the historical evolution of AI in medicine, where early rule-based systems like INTERNIST-I^42^ and MYCIN^43^ attempted to combine knowledge representation and reasoning for differential diagnosis. When the LLM was used to identify sepsis based solely on clinical notes (referred to as COMPOSER-LLM*_baseline_*), its performance was lower compared to the COMPOSER-LLM pipeline. This aligns with previous studies that have also noted the difficulties of using standalone LLMs for diagnostic purposes^44–46^, and highlights the importance of combining the strengths of predictive modeling (for initial risk stratification) and differential diagnosis (for addressing prediction uncertainty). Furthermore, incorporating differential diagnosis into predictive modeling can enhance patient safety by mitigating anchoring bias associated with false alarms focused on a single diagnosis. By systematically considering alternative diagnoses, clinicians can avoid premature closure and improve diagnostic accuracy. Similarly, in scenarios with high prediction uncertainty, differential diagnosis can mitigate the potential harm of automation bias (missed detections) by improving model sensitivity via a ranked list of potential diagnoses (top-k).

False alarms can often lead to alert fatigue, increase of physician’s cognitive burden, and can result in exposure of patients to unnecessary treatments^47^. Although in recent years adoption of Sepsis-3 consensus criteria for tagging patients for sepsis has resulted in significant progress in sepsis epidemiological studies and algorithm development^48^, such silver-standard methods have limited sensitivity and specificity^49^ and often further chart-review is warranted. In this study, two study authors performed an in depth analysis of a sample of 50 false-positive patients and were asked to identify if there was a suspicion of bacterial infection at the time of the alert. The review, which had significant agreement, revealed that 62% of the false positives had bacterial infections when the alert was triggered, indicating the potential clinical utility of the alert even for patients who did not progress to sepsis. Thus, even if sepsis was not present, a concern for a bacterial infection would likely have benefited from provider evaluation and intravenous antibiotics, which may have prevented development of sepsis.

Some of the notable features of the COMPOSER-LLM system include: 1) The use of the open-source Mixtral 8×7B Large Language Model in this study allowed us to deploy the entire COMPOSER-LLM pipeline within UCSD’s Health Insurance Portability and Accountability Act (HIPAA) compliant cloud environment. This deployment ensured that patient data remained securely within the hospital’s firewall, maintaining the protection of UCSD patient information; 2) The cloud-based real-time platform for COMPOSER-LLM was designed to be both scalable and flexible, enabling it to support real-time inference for models of any size. Consequently, the Mixtral 8×7B model, comprising 45 billion parameters, was used for real-time inference without encountering any computational bottlenecks; 3) The LLM-based clinical sign or symptom extractor was designed to provide explanation for each of the clinical signs or symptoms extracted, enhancing transparency and explainability for the clinical end-user. This provided an advantage over traditional NLP-based information retrieval techniques, which typically lack interpretability; 4) The decision to run the LLM-based differential diagnosis tool only for samples with scores between 0.5 and 0.75 enabled us to significantly save on computational costs as the multi-GPU instance hosting the LLM was intermittently switched on/off as and when required (on average 10 calls to the LLM microservice/day).

This work has several limitations. The differential diagnosis tool is currently triggered only after the availability of certain clinical notes (“ED provider note”, “History and Physical note”, “Diagnostic image report” or “Progress note”), potentially delaying alert generation among patients in the uncertainty interval of 0.5-0.75. However, when COMPOSER-LLM was deployed prospectively, the differential diagnosis tool was triggered even if a note was incomplete, as the contextual information within the notes could still be useful. Future research could explore using LLM-based queries for key patient and provider information (e.g., suspicion of infection) and real-time capture of provider notes through speech recognition and transcription to address missing or incomplete notes. Additionally, while a pre-trained LLM was used in this study, a fine-tuned LLM might further enhance performance by capturing more domain-specific knowledge. While fine-tuning an LLM with local data may enhance performance, it can also reduce generalizability across different hospital settings and pose practical challenges for widespread adoption. Our approach prioritizes the use of an off-the-shelf, smaller LLM to ensure broad applicability, particularly in resource-constrained environments.

In conclusion, our findings underscore the potential of LLMs in augmenting traditional prediction models for sepsis, particularly in high-uncertainty prediction scenarios. By effectively processing and analyzing unstructured clinical narratives, COMPOSER-LLM offers a promising advancement in predictive healthcare analytics. Future prospective studies are needed to assess how this approach impacts patient care and outcomes. Furthermore, LLM-based disease likelihood tools could help derive differential diagnoses^50^, improving the recognition of other life-threatening conditions that resemble sepsis.

## Methods

### Study cohort

Deidentified data from the EHR (using FHIR and HL7v2 standards) of patient encounters admitted to the Emergency Department (ED) within two University of California San Diego (UCSD) Health hospitals was used in this study. Patients aged 18 years or older were monitored throughout their stay until they experienced their first episode of sepsis, transitioned to comfort care measures, or were transferred out of the Emergency Department, whichever occurred earlier. To ensure a sufficient quantity of predictor data, we focused on sequential hourly predictions of sepsis starting two hours after ED triage. While the decision threshold of the previously described COMPOSER model^15^ was set to 0.6, the COMPOSER-LLM adopted a lower decision threshold of 0.5 to improve sensitivity. To address the potential increase in false alarm rate, the proposed model leveraged additional contextual information from clinical notes for the high-uncertainty predictions within the 0.5-0.75 range. To evaluate the effects of adjusting the prediction model to be more sensitive by lowering its decision threshold, and to explore the benefits of using an LLM to handle uncertain predictions, all patients with at least one COMPOSER risk score exceeding 0.5 were included for further analysis. Patients identified as having sepsis before the prediction start-time or those without heart rate or blood pressure measurements prior to this time were excluded. Predictions were made if the following criteria were met: 1) At least one vital and lab measurement in the past 24 hours; 2) No antibiotics received; and 3) Availability of “ED provider note”, “History and Physical note”, “Diagnostic Image report” or “Progress note”. The History and Physical (H&P) note is usually completed during a patient’s initial visit, and includes the patient’s medical history, family history, social history, and a review of systems. It also documents the physical exam findings and an initial assessment and plan. The ED provider note is created during an emergency room visit, and typically includes detailed documentation to capture the patient’s initial presentation, evaluation, treatment, and discharge or admission plan. Progress notes are daily or periodic updates documenting a patient’s progress, often using the SOAP format (Subjective, Objective, Assessment, Plan). They track changes in the patient’s condition, responses to treatment, and any new findings. A Diagnostic Image report contains summaries of imaging studies, like X-rays, CT scans, and MRIs, detailing the findings and the radiologist’s interpretation.

The overall retrospective dataset was divided into a *development cohort* and a *validation cohort*. The *retrospective development cohort* included patient encounters from September 2023, while the *retrospective validation cohort* comprised encounters from October to December 2023. Additionally, the COMPOSER-LLM pipeline was prospectively deployed in silent mode for real-time sepsis prediction in the two EDs within the UCSD Health system starting May 1, 2024. The prospective data collected during the time period of May 1 - June 15 2024 will be referred to as *prospective cohort*.

Patients were identified as having sepsis according to the Sepsis-3 international consensus definition for sepsis^2,51^. The onset time of sepsis was established by following previously published methodology, using evidence of organ dysfunction and suspicion of clinical infection^15,52,53^. Clinical suspicion of infection was defined by a blood culture draw and at least 4 days of non-prophylactic intravenous antibiotic therapy satisfying either of the following conditions: (1) if a blood culture draw was ordered first, then an antibiotic order had to occur within the following 72 h, or (2) if an antibiotic order occurred first, then a blood culture draw had to occur within the next 24 h. Evidence of organ dysfunction was defined as an increase in the Sequential Organ Failure Assessment (SOFA) score by two or more points. In particular, evidence of organ dysfunction occurring 48 hours before to 24 hours after the time of suspected infection was considered, as suggested in Seymour et al.^51^. This investigation was conducted according to University of California San Diego IRB approved protocol #805726 with a waiver of informed consent.

### COMPOSER model

COMPOSER is a previously published model that was trained to predict the onset of sepsis up to four hours in advance^54^. It was shown that the deployment of COMPOSER at two EDs within the UCSD Health system was associated with a 17% relative reduction in in-hospital sepsis mortality as part of a quality improvement initiative in our EDs^15^. This deep neural network model integrates routinely collected laboratory and vital signs, along with patient demographics (age and sex), comorbidities, and medications, to generate a risk score for sepsis onset within the next four hours. Additionally, the model utilizes a conformal prediction module to reject out-of-distribution samples that may arise due to data entry errors or unfamiliar cases.

### Differential diagnosis tool

For samples with COMPOSER risk scores within a high uncertainty interval, an LLM-based differential diagnosis tool was employed to enhance diagnostic certainty. This high uncertainty interval was characterized by a low and nearly constant positive predictive value (see Supplementary Figure 1). In this study, differential diagnoses were conducted for a total of 19 conditions, including severe sepsis and sepsis-mimics (i.e., cardiogenic shock, pulmonary embolism, hypovolemia, hypovolemic shock, cirrhosis, heart failure exacerbation, alcohol/drug withdrawal, gastrointestinal hemorrhage, anaphylaxis, acute respiratory distress syndrome, influenza, covid-19, myocardial infarction, coronary artery disease, malignancy, pancreatitis, heat stroke and diabetic ketoacidosis). The tool aimed to increase diagnostic certainty by confirming the presence of clinical signs and symptoms related to severe sepsis and its mimics in the clinical notes. It used an LLM to first extract these relevant signs or symptoms, which were then analyzed using likelihood calculators to estimate the probability of each differential. The generated list, ranked by likelihood, was then used downstream to determine if an alert was to be fired or not.

### LLM-based clinical sign or symptom extractor

We utilized the instruction fine-tuned Mixtral 8×7B LLM^55^ to extract clinical signs and symptoms from unstructured clinical notes. The LLM-based data extraction pipeline was designed to accept a prompt and clinical notes (all notes generated from admission to the time of prediction) as input and produce a text output in JSON format. The schematic diagram of the LLM-based data extraction pipeline is shown in Figure 1.

Because the length of clinical notes can exceed the predefined input length (context size) of the LLM, we employed the retrieval augmented generation (RAG) technique to extract smaller, relevant text chunks (context) for the queried clinical sign or symptom. These extracted text chunks were then appended to the input prompt of the LLM^28^. The RAG pipeline was implemented as follows: 1) Clinical notes generated from the time of admission until the prediction were segmented into smaller text chunks; 2) These segmented text chunks were passed through an embedding model to obtain embedding vectors; 3) The embedding vectors were stored in a vector database; 4) The embedding vector corresponding to the query text was extracted (query embedding); and 5) The top ‘*K’* embedding vectors, most similar to the query embedding vector, were retrieved from the vector store, and the corresponding text chunks were appended to the input prompt of the LLM.

The RAG pipeline used the Chroma vector database for storing and retrieving embedding vectors and HuggingFace’s Instructor model^56^ for converting text to embedding vectors. The text chunk size was set to 1000 tokens with a 300 token overlap. The total number of embedding vectors to be retrieved from the vector store was set to five (*K*=5).

The input prompt was designed to instruct the LLM to review the patient’s medical note and identify the presence of the queried clinical sign or symptom. The prompt also provided instructions to the LLM to generate the output in a JSON format. The prompt used in our analysis was as follows:

*“You are an ED doctor. Your task is to identify the following abnormal clinical signs and symptoms: {clinical sign or symptom}. Think step-by-step and provide your response in the following JSON format: {<CLINICAL sign or symptom> : [“Yes or No”, “Concise justification?”]} Medical note: {RAG context}.”*

To minimize hallucinations and to maintain consistency in text generation, the temperature parameter of the LLM was set to 0.3 (See Supplementary Note 1 for more details). Additionally, for each clinical sign or symptom, the LLM pipeline was run three times and the majority outcome (clinical sign or symptom present or not) across the multiple runs was used for downstream tasks. Finally, the entire LLM-based clinical sign or symptom extraction pipeline was implemented using the LangChain framework in Python.

### Clinical sign or symptom-based disease likelihood calculator

A Bayesian likelihood calculator was used to compute the likelihood of a disease based on the clinical signs or symptoms identified by the LLM. The posterior probability of a disease given a set of clinical signs or symptoms 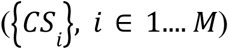, *P*(*D*|*CS*), was calculated as follows:

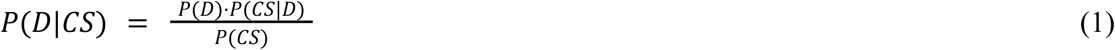

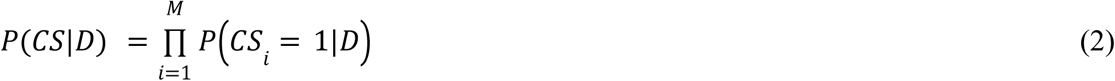

Where, *CS*_*i*_ = 1 corresponded to the scenario under which the clinical signs or symptom *CS*_*i*_ was identified to be present by the LLM pipeline. *M* corresponded to the total number of clinical signs or symptoms defined for a given disease. D refers to the various sepsis-mimics considered in this study. Please see the subsection *“Differential diagnosis tool”* for a list of all the sepsis-mimics.

The clinical signs or symptoms for severe sepsis and the sepsis-mimics were derived with the help of two of the study authors (G.W and J.C.A). The final set of clinical signs or symptoms used in this study were: elevated inflammatory markers, positive influenza test, altered mental status, dehydration + sweating, metabolic acidosis, abdominal pain, ketonuria, elevated liver enzymes, ascites, risk factors for lung injury, urticaria + angioedema, seizures, history of substance abuse, hypoxemia, jaundice, positive biopsy or imaging, risk factors for thrombosis, tachypnea, cough + sore throat, exposure to heat, elevated BNP, elevated tryptase + histamine, myalgia + headache, pulmonary edema, elevated lactate, elevated glucose, fever, elevated cardiac enzymes, positive covid-19 test, hematemesis or melena, dry mucous membranes, bilateral infiltrates on chest x-ray, suspicion of bacterial infection, elevated d-dimer, low urine output, tachycardia, positive toxicology screen, nausea + vomiting, cough + dyspnea, history of fluid loss, anemia, elevated lipase, positive blood culture, exposure to allergens, loss of taste or smell, ECG changes, hypotension, organ dysfunction, chest pain, weight loss + fatigue, dyspnea.

The sorted list of differentials (based on their likelihood) was then used downstream to determine if an alert was to be fired or not. Additionally, the likelihood values of clinical signs or symptoms conditioned on each of the diseases (*P*(*CS*_*i*_ = 1|*D*)) were optimized using Bayesian optimization on the retrospective development cohort.

### COMPOSER-LLM

The schematic diagram of the entire COMPOSER-LLM pipeline is shown in Figure 1. Starting from the time of ED admission, COMPOSER generated a sepsis risk score at an hourly resolution. If the risk score was greater than a primary decision threshold (θ_1_ =0.75) a notification was sent via a nurse-facing best practice advisory. Risk scores closer to a secondary decision threshold (θ_2_ =0.50) were often associated with false alarms. Therefore, for risk scores within the high-uncertainty region (θ_2_ ≥ risk score > θ_1_), the LLM-based differential diagnosis tool was utilized to improve diagnostic certainty for sepsis. Specifically, if ‘Severe Sepsis’ was present within the top-5 sorted differentials and the LLM identified a ‘suspicion of bacterial infection’, a notification was sent via a nurse-facing Best Practice Advisory.

Please note that throughout this paper, COMPOSER-LLM and COMPOSER-LLM*_DDx_* refer to the same pipeline, unless otherwise noted.

### Baseline model comparison

Instead of extracting the clinical signs and symptoms followed by performing differential diagnosis (COMPOSER-LLM*_DDx_*), we explored the possibility of directly asking the LLM to identify the presence of severe sepsis in a patient based on clinical notes (which we’ll refer to as COMPOSER-LLM*_baseline_*). In this experiment, the LLM generated a binary output for “Sepsis”. The prompt used was as follows:

*“You are an ED doctor. Your task is to identify if sepsis is present in the current admission. Think step-by-step and provide your response in the following JSON format: {“ Sepsis” : [“Yes or No”, “Concise justification?”]}*

*Note that “Sepsis” is defined as suspicion or documentation of infection with evidence of organ dysfunction*.

*Medical note: {RAG context}.”*

For samples with COMPOSER risk scores between 0.5 and 0.75 (θ_2_ = 0.5, θ_1_ = 0.75), an alert was fired if the output from the LLM for “Sepsis” was “Yes”. Note that, for COMPOSER risk scores greater than the primary decision threshold (θ_1_), an alert was fired irrespective of the output from LLM.

An additional baseline model considered in this study was the COMPOSER-LLM*_SLT_* pipeline. COMPOSER-LLM*_SLT_* utilized a sepsis likelihood tool to calculate the likelihood of severe sepsis based on the presence of severe sepsis related clinical signs or symptoms. For samples with COMPOSER risk scores between 0.5 and 0.75 (θ_2_ = 0.5, θ_1_ = 0.75), an alert was fired if the sepsis likelihood score exceeded a predetermined likelihood-based decision threshold (α) and the LLM identified a ‘suspicion of bacterial infection’. Note that, for COMPOSER risk scores greater than the primary decision threshold (θ_1_), an alert was fired irrespective of the output from LLM.

### Experimental setup and evaluation

For all continuous variables, we have reported medians ([25th–75th percentile]). For binary variables, we have reported percentages. Differences between the septic and non-septic cohort were assessed with Wilcoxon rank sum tests on continuous variables and Pearson’s chi-squared tests on categorical variables and significance was assessed at a p-value of 0.05. For binary variables, we have reported percentages. Sensitivity, positive predictive value (PPV), and F-1 score at a fixed decision threshold have been reported at the encounter level. Additionally, the following policy was utilized for COMPOSER-LLM alerts: 1) an alert was considered a true positive if ‘Severe Sepsis’ was present within the Top-5 differentials for a septic patient, 2) an alert was considered a false positive if ‘Severe Sepsis’ was present within the Top-1 differentials for a non-septic patient.

COMPOSER-LLM was designed as a notification-only tool to predict onset time of sepsis four hours in advance, no earlier than 48 hours in advance and under a silencing policy of six hours. Specifically, Sensitivity, PPV and F-1 score were reported under an end-user clinical response policy in which alarms fired up to 48 hours prior to onset of sepsis were considered as true alarms, and the model was silenced for six hours after an alarm was fired. The end-user clinical response policy is identical to the policy described previously in our published work^15,54^. Additionally, we have reported false alarms per patient hour (FAPH) which can be used to calculate the expected number of false alarms per unit of time in a typical care unit (e.g., a FAPH of 0.025 translates to roughly 1 alarm every 2 h in a 20-bed care unit). The FAPH was calculated by dividing the total number of false alarms by the total number of data points (sum of hourly time points across all patients) in a given cohort.

All hyperparameters of the LLM pipeline including text chunk size (1000 tokens), number of retrieval vectors from vector database (*‘K’=5*), temperature of the LLM (0.3), likelihood values of clinical signs or symptoms conditioned on sepsis were optimized using Bayesian hyperparameter optimization on the retrospective development cohort.

The COMPOSER model was implemented in TensorFlow (version 2.13.0). For the LLM pipeline, we utilized the instruction fine-tuned Mixtral 8×7B LLM model (version 0.1, GPTQ quantized) (HuggingFace link: https://huggingface.co/TheBloke/Mixtral-8x7B-Instruct-v0.1-GPTQ). The LLM-based clinical signs or symptom extraction pipeline was implemented using the LangChain framework (version 0.1.5) in Python (version 3.11.5). The LLM pipeline was run on an AWS multi-GPU EC2 instance with NVIDIA A10G GPUs (ec2 instance type: g5.12xlarge).

### Clinician chart review

The goal of utilizing the LLM-based differential diagnosis tool was to reduce the false alarm rate of the standalone COMPOSER pipeline while maintaining similar sensitivity levels. To further investigate the false positives generated by the COMPOSER-LLM model, a clinician chart review was conducted. Chart review of the false positives was performed by two physicians and study authors (J.C.A and A.P). Each physician was asked to review the chart of a given patient and identify if there was a ‘suspicion of bacterial infection’ at the time of alert. In cases of disagreement between the two clinicians, a third physician and study author (G.W) provided the final evaluation.

### COMPOSER-LLM prospective deployment

The COMPOSER-LLM model was prospectively deployed in silent-mode on a cloud-based platform, as previously described by Boussina et al.^15^. Prospective validation studies are essential in clinical applications of LLMs as retrospective performance may not accurately reflect real-world performance due to factors such as incomplete or missing clinical notes. The real-time platform extracted data at an hourly resolution of all the active patients (across the two Emergency Departments within UCSD Health system) using FHIR APIs with OAuth 2.0 authentication, and passed the input feature set (consisting of laboratory measurements, vitals measurements, comorbidities, medications, clinical notes, and demographics) to the COMPOSER-LLM inference engine. The inference engine consisted of COMPOSER microservice and differential diagnosis tool microservice hosted within separate EC2 instances. The sepsis risk scores generated by the COMPOSER-LLM pipeline were then written to a flowsheet within the EHR using an HL7v2 outbound message. The flowsheet then triggered a nurse-facing Best Practice Advisory (BPA) that alerted the caregiver that the patient was at risk of developing severe sepsis. As COMPOSER-LLM was deployed in silent mode, the BPA was not shown to the end-user. The schematic diagram of the real-time deployment pipeline is shown in Figure 2. The COMPOSER-LLM pipeline was deployed for real-time prediction of sepsis across the two EDs within the UCSD Health system starting from May 1, 2024.

## Data availability

Access to the de-identified UCSD cohort can be made available by contacting the corresponding author and via approval from the UCSD Institutional Review Boards (IRB) and Health Data Oversight Committee (HDOC).

## Code availability

The code for computing performance metrics is available at https://github.com/NematiLab/COMPOSER-LLM.

## Acknowledgements

S.N. is funded by the National Institutes of Health (#R01LM013998, #R35GM143121, #R42AI177108). G.W. has been supported by the National Institutes of Health (#K23GM146092). This work has been supported by the National Institutes of Health (#4R42AI177108-01). K.S.’s institution is currently supported by funding from the National Institutes of Health for unrelated work; his institution previously received grant funding from Teva Pharmaceuticals for unrelated work; and he previously consulted for Flatiron Health for unrelated work. The opinions or assertions contained herein are the private ones of the author and are not to be construed as official or reflecting the views of the NIH or any other agency of the US Government.

## Author contributions

S.P.S., E.A.S and S.N. were involved in the original conception and design of the work. S.P.S and S.N. developed the network architectures, conducted the experiments, and analyzed the data. S.M assisted with analyzing the data. R.K assisted with setting up the computational environment and conducted experiments. A.P, G.W., J.C.A, K.S and E.A.S. provided clinical expertise, reviewed patient data, and contributed to interpretation of results. All authors contributed to manuscript preparation, critical revisions, and have read and approved the manuscript.

